# Age-related differences in psychopathology within sex chromosome trisomies

**DOI:** 10.1101/2024.11.22.24317803

**Authors:** Melissa R. Roybal, Siyuan Liu, Isabella G. Larsen, Anastasia Wass, Lukas Schaffer, Tiffany Ajumobi, Ethan T. Whitman, Allysa Warling, Liv Clasen, Jonathan Blumenthal, Srishti Rau, Armin Raznahan

**Affiliations:** National Institutes of Health, National Institute of Mental Health, Bethesda, MD, United States; Georgetown University School of Medicine, Georgetown, Washington, D.C., United States; Institute for Behavioral Genetics, University of Colorado Boulder, Boulder, CO, United States; Department of Psychology and Neuroscience, University of Colorado Boulder, Boulder, CO, United States; School of Medicine, The Johns Hopkins University, Baltimore, MD, United States; Department of Psychology & Neuroscience, Duke University, Durham, NC, United States; Harvard Medical School, Boston, MA, United States; Children’s National Health System, Center for Autism Spectrum Disorders and Division of Neuropsychology, Washington, D.C., United States

**Keywords:** sex chromosomes, sex chromosome trisomy, neurogenetic conditions, psychopathology, developmental psychopathology

## Abstract

Sex chromosome trisomies (SCTs) are a group of genetic disorders characterized by presence of a supernumerary sex chromosome, resulting in karyotypes other than XX or XY. These include XXX (Trisomy X), XXY (Klinefelter syndrome), and XYY (Jacobs syndrome). Sex chromosome trisomies have been linked to increased risk for psychopathology; however, this relationship warrants additional research. Specifically, little is known regarding potential age-related variation in risk for psychopathology and how this may differ across karyotypes and subdomains of psychopathology, which has relevance for psychoeducation, personalized care, and mechanistic research. Thus, we used the Child Behavior Checklist (CBCL) to estimate age-related variation in psychopathology in a large cross-sectional sample of individuals with SCTs (n = 201) and euploidic controls (n = 304) spanning the age range of 5-18 years. We found that elevations of psychopathology in SCT were significantly associated with age in a manner that varied as a combined function of the karyotype and CBCL scale being considered. Post hoc tests revealed there is a uniquely pronounced age-associated increase in severity of social problems in XYY, alongside a lack of statistical evidence for age-related variation in the severity of psychopathology for other CBCL domains and SCT karyotypes. Our findings are relevant for advancing the personalization of clinical assessment and monitoring in SCTs. They also highlight potential windows of dynamic risk emergence for closer clinical and biological study, as well as opportunities to provide intervention to mitigate future risk.

## INTRODUCTION

Sex chromosome trisomies (SCTs) are a group of genetic disorders characterized by presence of a supernumerary sex chromosome, resulting in karyotypes other than XX or XY. These include XXX (Trisomy X), XXY (Klinefelter syndrome), and XYY (Jacobs syndrome). These conditions are individually rare, but collectively common. XXX occurs in 1 per 1000 newborn females, XXY and XYY in 1.5 and 1.1 per 1000 newborn males respectively, with a combined prevalence across karyotypes of ∼ 1 in 500 newborns [1]. All 3 SCTs have been associated with increased risk for cognitive impairment and psychopathology (i.e., social-emotional and behavioral difficulties that may or may not reach clinically elevated or diagnostic thresholds). Studies in both clinically ascertained and population-based SCT cohorts have reported elevated rates of various neuropsychiatric symptoms which often reach clinical thresholds for psychiatric diagnoses. These include symptoms associated with autism spectrum disorder (ASD), attention deficit hyperactivity disorder (ADHD), anxiety disorders, and mood disorders [2-9]. Despite the fact that all SCTs increase risk for multiple domains of psychopathology, there is some evidence that the specific profile of risk may vary between different SCT karyotypes. For example, there is a tendency towards relatively higher social and externalizing impairments in XYY, and relatively higher depressive problems in XXY and XXX [3, 10-12]. These prior findings highlight the importance of studying SCTs individually and in comparison to each other, rather than collapsing them into a monolithic group for comparison with euploidic controls.

While all SCTs increase risk for psychopathology, very little is known regarding potential variation in this risk over development. It is certainly clear that rates of psychopathology vary dramatically with age in the general population [13-15] and in psychiatrically diagnosed populations [16-17], highlighting the potential that SCT effects on psychopathology may also vary with age. To date, one of the only available studies of age-related variation of psychopathology in SCT focused on affective symptoms as well as oppositional defiant disorder (ODD) and ASD-related features during the preschool years [18]. Researchers found that affective, pervasive developmental, and oppositional defiant problems appeared to worsen with age in a combined group of all 3 SCT karyotypes [18]. However, we currently lack a survey of age-related variation in psychopathology that compares different SCT karyotypes across a broader developmental window and a broader range of psychopathology. Consequently, it remains unclear if there is any age-related variation in risk for psychopathology in SCT and whether any such variation differs as a function of SCT karyotype and/or the specific domain of psychopathology in question. Addressing these gaps in knowledge would support better personalization of clinical assessment and monitoring in SCT and highlight potential windows of dynamic risk emergence for close clinical and biological study as well as intervention to mitigate future risk.

Given the above considerations, the current study systematically profiles scores for 11 complementary dimensional measures of psychopathology from the commonly used Child Behavior Checklist (CBCL) [19, 20] in a large cross-sectional sample of individuals with SCT and controls spanning the age range of 5-18 years. We estimate age-related variation in psychopathology for each sampled karyotype and CBCL dimension, enabling us to test for instances of statistically and clinically significant dynamism of psychopathology across age in SCT. Given the absence of prior work examining this question, our analyses provide a valuable initial screen for the presence and nature of age-varying psychopathology in SCTs.

## METHODS

### Participants

The present study includes data from a total of 505 individuals with a mean age of 12.2 years (range 5-18 years). This sample includes 201 individuals with SCTs (XXX = 50, XXY = 92, XYY = 59) and 304 euploidic volunteers (XX = 151, XY = 153). Sample demographics are shown in **Table 1**.

**Table 1.**
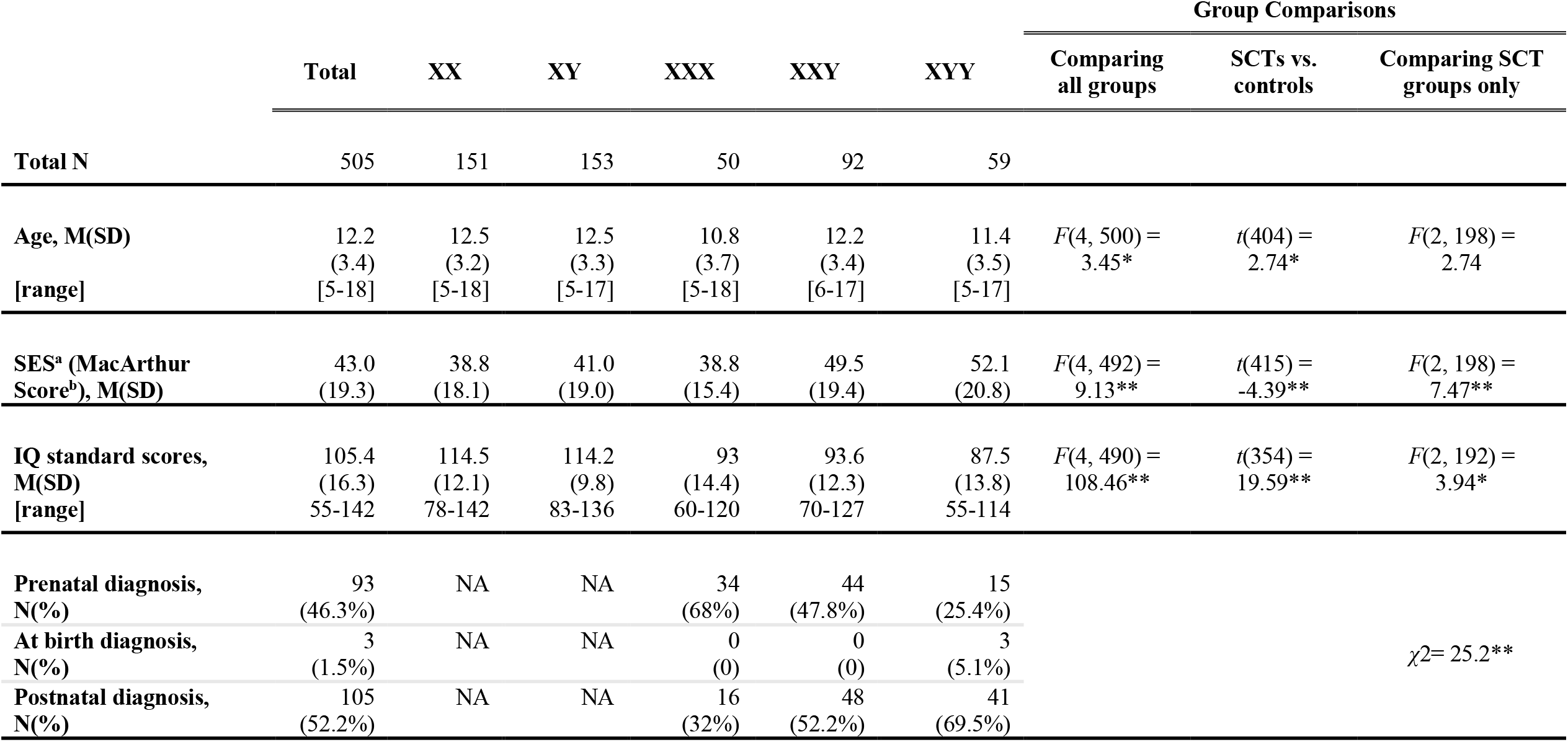
Sample demographics. * *p* < .05, \*\**p* < .001 ^a^Socioeconomic status, ^b^The MacArthur Scale of Subjective Social Status [21]

Participants with SCTs were recruited in collaboration with the Association for X and Y Chromosome Variations (AXYS; genetic.org) and the National Institutes of Health (NIH) Clinical Center Office of Patient Recruitment – the latter of which also assisted in the recruitment of XX and XY controls. All XX and XY controls were screened using a standardized interview to verify the absence of a prior psychiatric diagnosis or early developmental difficulties requiring provision of extra support at school or home. Inclusion criteria for the SCT groups were (i) a non-mosaic SCT diagnosis that was confirmed by karyotype analyses or review of prior medical records for those SCT participants unable to give blood, (ii) no history of brain injury or comorbid neurological disorder (e.g., epilepsy), and (iii) age between 5 and 25 years on the day of assessment. All research assessments were conducted at the NIH Clinical Center, Bethesda, MD, USA. Written informed consent was secured from adult participants and parents of minor participants and written assent from children. The study was approved by the National Institute of Mental Health (NIMH) Institutional Review Board (Clinical trial reg no. NCT 00001246, clinicaltrials.gov; NIH Annual Report Number, ZIA MH002794-13; Protocol number: 89-M-0006).

### Measurement of psychopathology and cognitive ability

We measured psychopathology in all participants using the caregiver-completed CBCL [19, 20]. Two versions of this measure were used, partially due to differences in the age ranges that they cover. The earlier CBCL [19] is approved for children as young as four while the later version [20] begins at age six. This commonly used instrument combines item-level responses to derive eight syndrome scales (Anxious/Depressed, Withdrawn/Depressed, Somatic Complaints, Social Problems, Thought Problems, Attention Problems, Rule-Breaking Behavior, and Aggressive Behavior). Three composite broadband scales are also derived: Externalizing Domain (Rule-Breaking Behavior and Aggressive Behavior), Internalizing Domain (Anxious/Depressed, Withdrawn/Depressed, and Somatic Complaints), and Total Problems (includes items from all syndrome scales plus an additional 17 items that do not belong to any specific syndrome scale). Ratings on each scale are expressed as T-scores (M=50; SD=10) derived based on comparisons to published normative data stratified by age and gender.Higher scores indicate the presence of more problem behaviors/impairment. For the eight syndrome scales, T-scores of 65-69 are borderline clinical, and T-scores of 70 or higher are in the clinical range; for the three broadband scales, T-scores of 60-63 are borderline clinical, and T-scores of 64 or higher are clinically elevated. By focusing our analyses of age-related effects on T-scores (see below), levels of psychopathology in all individuals are norm-standardized to the same set of reference distributions within the CBCL. Initially, caregivers completed this measure using pencil and paper. However, during the COVID-19 pandemic we shifted to online administration of the CBCL via REDCap (Research Electronic Data Capture), an electronic data capture tool hosted at National Institute of Mental Health [22, 23]. REDCap is a secure, web-based software platform designed to support data capture for research studies. Berg et al. [24] indicated there are no significant differences in CBCL results whether completed on paper or on a computer. Thus, we combined data from all sources of administration (paper/pencil and computer) for these analyses.

General cognitive ability (henceforth Intelligence Quotient “IQ”) was measured for all participants using Weschler scales. For the SCT groups, Full Scale IQ was estimated from the Wechsler Preschool and Primary Scale of Intelligence, Third Edition (WPPSI-III) [25], Wechsler Preschool and Primary Scale of Intelligence, Fourth Edition (WPPSI-IV) [26], Wechsler Intelligence Scale for Children, Fifth Edition (WISC-V) [27], Wechsler Adult Intelligence Scale, Fourth Edition (WAIS-IV) [28], Wechsler Abbreviated Scale of Intelligence (WASI), First Edition [29], or the Wechsler Abbreviated Scale Intelligence, Second Edition (WASI-II) [30]. For the euploidic controls, IQ was measured using the WASI, WASI-II, Wechsler Intelligence Scale for Children, Third Edition (WISC-III) [31], Wechsler Intelligence Scale for Children-Revised (WISC-R) [32], Wechsler Adult Intelligence Scale-Revised (WAIS-R) [33], or for 25 XY males, vocabulary and matrix reasoning from the WASI-II were administered over telehealth. Two controls received alternative IQ tests, specifically the Differential Abilities Scale, 2nd Edition (DAS-II) [34] and the Kaufman Brief Intelligence Test, 2nd Edition (KBIT-2) [35]. IQ ratings are expressed as a standard score (M=100; SD=15) in comparison to normative expectations based on age. Higher scores indicate higher intellectual functioning.

### Statistical Analysis

All statistical analyses and data visualizations were completed using the R language for statistical computing, version 4.3.1 [36]. CBCL scores and age were visualized using ggplot2 [37] and Superheat [38]. Age was included as a centered variable so that intercept coefficients refer to effects observed at the mean age.

We tested for the omnibus effect of 3-way interactions between age, karyotype and scale on CBCL T-scores using an F-test for the ß_7_ term in the linear mixed model ([1]) below which includes a random effect to account for dependence of CBCL subscales within each person:

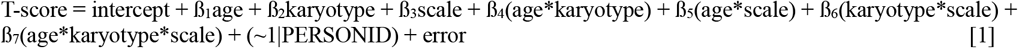

Presence of a significant 3-way interaction was then followed up with post hoc linear models examining the two-way interactions between (i) age and karyotype for each CBCL scale, and (ii) age and CBCL scale for each karyotype using ß_3_ coefficients from models [2] and [3] (respectively) below (and a nominal *p* < .05 cut off for statistical significance):

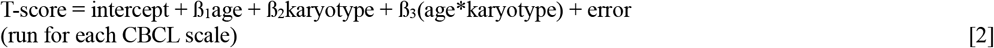

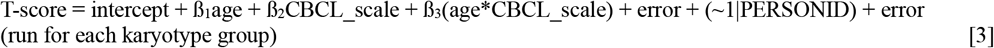

Tukey post hoc tests were run on outputs of model [2] to test for group differences in average CBCL T-score values at mean age for each scale.

Finally, linear effects of age in psychopathology within individual karyotype-scale combinations were estimated using the ß_1_ coefficient from linear model [4] below:

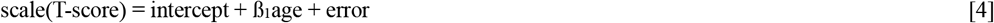

## RESULTS

### Participants

Participant characteristics are detailed in **Table 1** and **Supplementary Table 1**. As previously reported, karyotype groups differed by IQ and SES [10, 39].

We observed a statistically significant omnibus effect of group on age [*F*(4, 500) = 3.45 *p* = .009], although all groups spanned the same age range, and mean age per group remained within a narrow range of 10.8 to 12.5 years. Age did not vary in a statistically significant manner across the three SCT groups [*F*(2, 198) = 2.74 *p* = .07]. Of note, these subtle differences in age distribution are not transmitted to downstream analyses of psychopathology as (i) CBCL T-scores are age- and sex-standardized using the same set of published norms within the CBCL and (ii) our primary question of interest involves testing for age-related variations in these T-scores.

### Effects of age, karyotype and their interaction on psychopathology

Our study design mapped age-related variations in psychopathology for all combinations of five karyotype groups and 11 CBCL-derived measures (**Figure 1**).

**Fig. 1.**
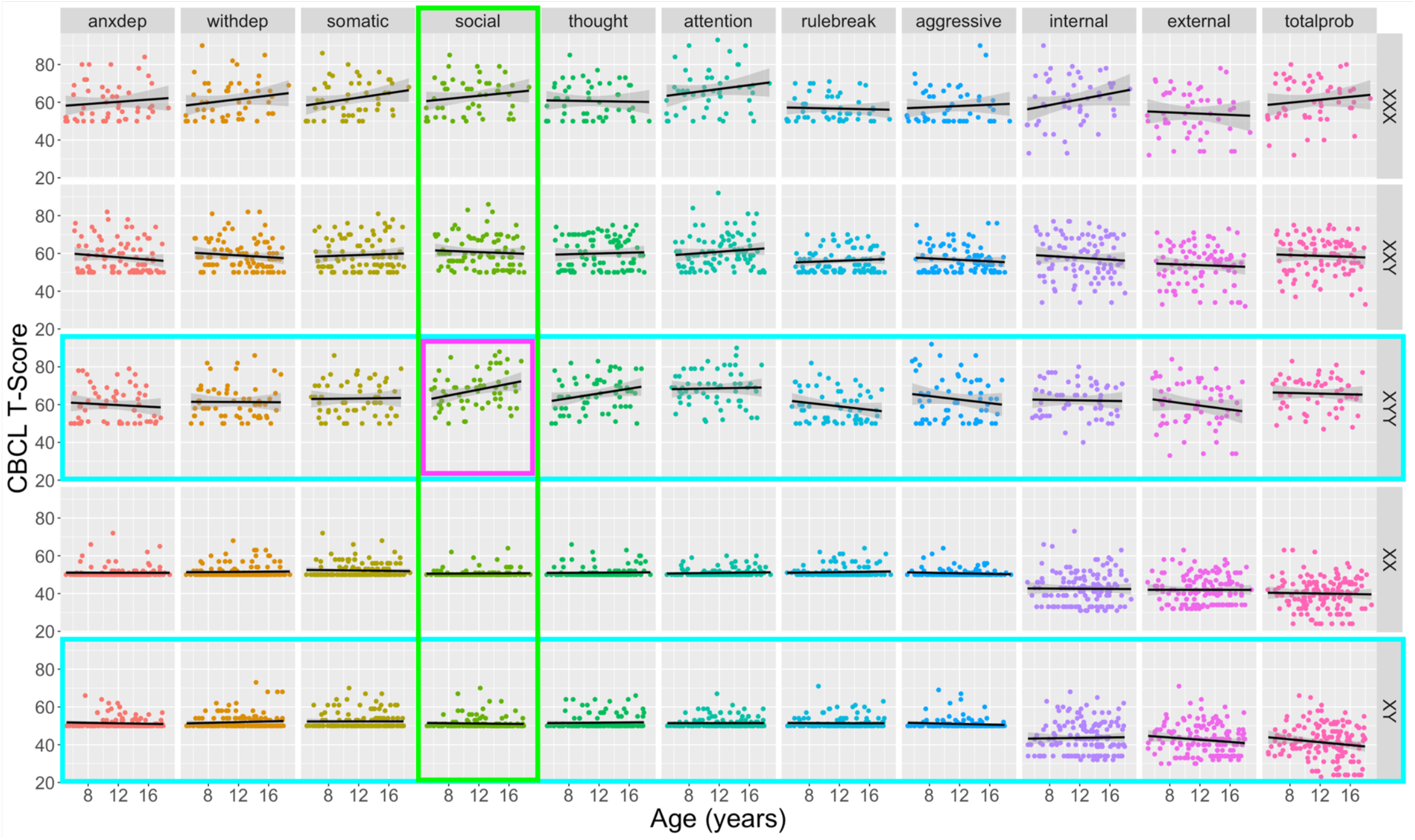
Scatterplots for distribution of individual-level psychopathology scores as a combined function of age and CBCL scale for all study karyotype groups. An omnibus F-test revealed a statistically significant 3-way interaction between age, scale, and karyotype in predicting CBCL T-scores [*F*(40, 4950) = 1.91, *p* = .0005]. Colored boxes represent significant (*p* < .05) follow-up post hoc tests for 2-way interactive effects of age with scale (columns, significance = green highlight) and karyotype (rows, significance = turquoise highlight) in prediction of CBCL T-scores. The purple panel outline highlights the specific scale-karyotype combination driving these significant post hoc tests: a prominent age-related increase of social problems in XYY syndrome that is absent for other karyotypes and psychopathology scales. Black fit lines show the ordinary least squares regression line for age prediction of T-scores for each karyotype-CBCL scale combination.

We observed a statistically significant 3-way interaction between age, karyotype and CBCL scale as predictors of T-score variation [*F*(40, 4950) = 1.91, *p* = .0005]. We therefore used post hoc analyses of the underpinning 2-way interactions to localize drivers of this 3-way interaction.

Testing the 2-way interaction between age and group for each CBCL scale revealed a significant karyotype-dependent difference in age effects on the CBCL Social Problems scale [*F*(4, 495) = 3.50, *p* = .008]. Data visualization and estimation of age effects on CBCL Social scale T-scores for each karyotype revealed that this interaction was driven by a uniquely prominent and statistically significant positive association between age and CBCL Social Problems T-scores in the XYY group [β = 0.78, *t*(57) = 2.27, *p* = .027] which was absent in all other karyotype groups. Reinforcing this finding, tests for the 2-way interaction between age and CBCL scale for each SCT karyotype group revealed significant scale-dependent differences in age effects on CBCL T-scores for the XYY group [*F*(10, 570) = 2.79, *p* = .002] – driven by the aforementioned age-related increase of Social Problems in XYY syndrome that was absent for other CBCL scales in XYY syndrome. We also observed a significant 2-way interaction between age and CBCL scale for prediction of T-scores in the XY karyotype [*F*(10, 1510) = 2.00, *p* = .030], indicating that age effects on CBCL T-score differ by scale in the XY group. However, age was not significantly associated with CBCL T-score for any individual scale in the XY group, although there was a statistically non-significant trend towards lessening Total Problems CBCL T-score with increasing age (β = -0.38, *p* = .093).

Taken together, these analyses indicate there is a uniquely pronounced age-associated increase in severity of social problems in XYY, alongside a lack of statistical evidence for differential age-related variation in other CBCL domains and SCT karyotypes. However, given substantial qualitative variation in the relationship between psychopathology and age across the full set of karyotype-scale combinations sampled in our study design (**Figure 1**), we provide a survey of age effects (i.e., impact of 1 year increase in age on CBCL T-scores as derived from model [4] above) on psychopathology for all unique scale-karyotype combinations represented in our study design (**Figure 2**). These age effects range from -0.53 to 0.78 (mean 0.03) across all observed karyotype-scale combinations in SCT. Although these linear age effects only reach statistical significance in post hoc testing for Social Problems in XYY (β = 0.78, *p* = 0.027), similarly sized age effects are also observed for Internalizing Problems in XXX. It is also notable that there is a marked, albeit not statistically significant, decrease in rule-breaking, aggressive, and overall externalizing behaviors in XYY syndrome. Providing important validation and context for this heterogeneity of age effects on psychopathology across karyotypes and CBCL scales in SCTs, we observe substantially weaker relationships between age and psychopathology for all CBCL scales in XY and XX groups. The mean absolute age effect across all CBCL scales in euploidic controls was 0.07 (SD = 0.09) as compared to a mean absolute age effect of 0.28 (SD= 0.21) across all CBCL scales in SCT groups – reflecting a tendency for greater age-related dynamism in CBCL scores in SCTs. We do however observe a trend towards an age-related lessening of Total Problems scores in XY males, apparently driven by lessening severity of Externalizing Problems (**Figure 2**).

**Fig. 2.**
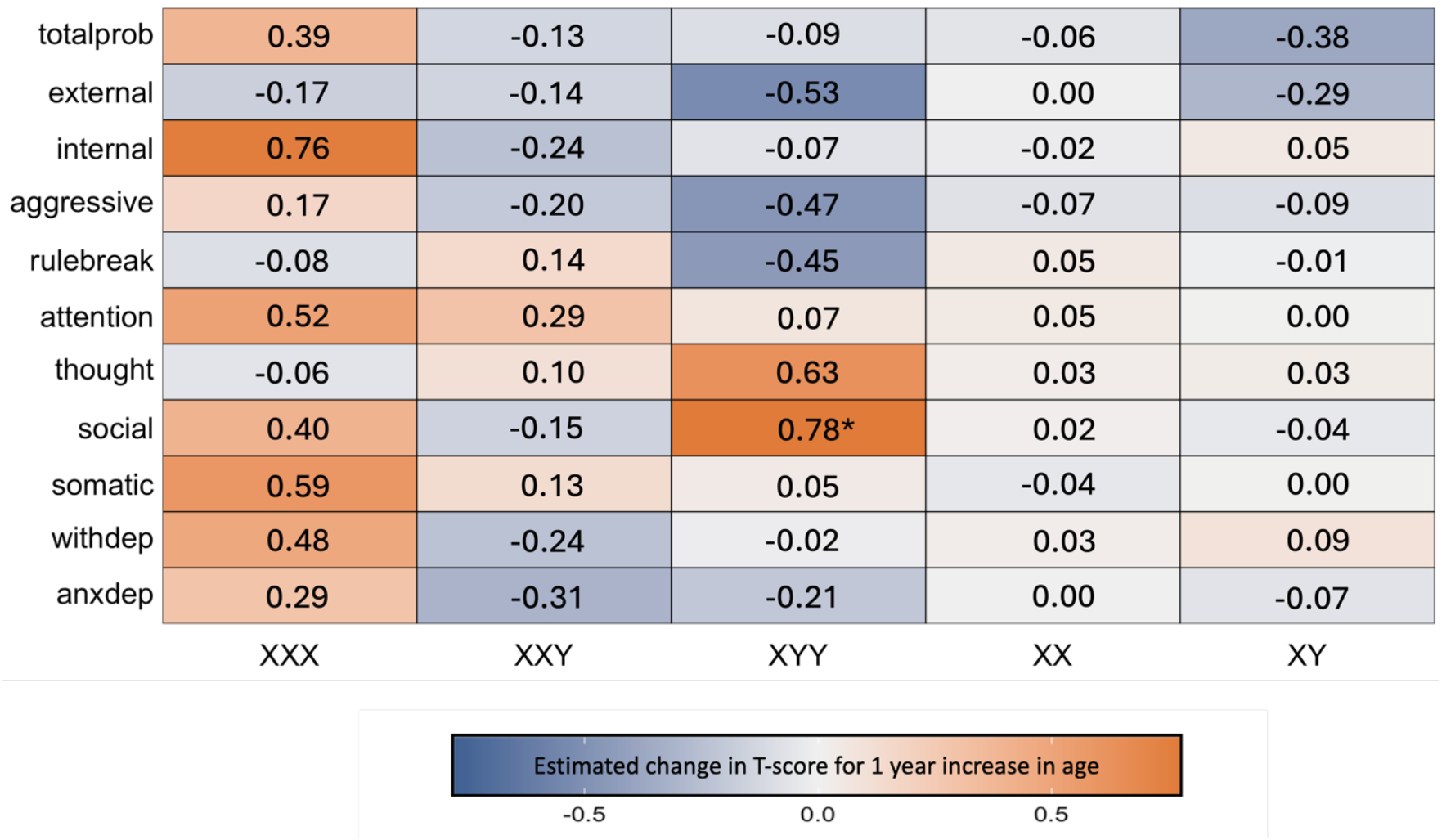
Effects of age on severity of psychopathology in all unique combinations of SCT karyotype and CBCL scales. This heatmap compiles linear coefficients for the effect of age on CBCL T-score (estimated mean CBCL T-score increase for one year increase in age) for all examined scale-karyotype combinations (*nominal *p* < .05).

The modeling approach required to address our primary goal of estimating age effects on psychopathology also yielded estimates of the mean effect of each SCT group on CBCL T-scores at the mean age of our sample. Notwithstanding age-related variations in CBCL T-scores (**Figures 1 and 2**), these main effect terms for each SCT group provide useful information regarding differences in the overall level of psychopathology across karyotypes. All SCT groups had statistically significant elevations of mean CBCL T-score relative to their gonadal controls for all CBCL domains (**Supplementary Table 2**). Inter-SCT group comparisons found a higher mean CBCL T-score in: (i) XYY as compared to both other SCTs for Social Problems, Thought Problems, and Aggressive Problems subscales as well as the Externalizing and Total Problems broadband scales, (ii) XYY as compared to XXY for Somatic Complaints, Rule-Breaking Behavior, and Internalizing, and (iii) XXX and XYY as compared to XXY for Attention Problems.

## DISCUSSION

Our study provides the first characterization of age-related variation in psychopathology within SCTs across childhood and adolescence and spanning multiple domains of symptom measurement. We consider four major insights of note below, before discussing study limitations and caveats, as well as implications for next steps.

First, surveying multiple domains of psychopathology in all three SCTs highlights a prominently positive association between age and social problems in XYY. Thus, not only do overall Social Problems (a scale which assesses one’s ability to navigate social situations) appear to be more severe in XYY than XXY or XXX groups (**Supplementary Table 2**), but they appear to also be uniquely exacerbated with increasing age in XYY. If replicated in future studies, this finding could potentially inform clinical practice by advising especially close monitoring of social problems over development in XYY syndrome. A confirmed trajectory of worsening social impairments would further strengthen the already strong case for promoting early detection and treatment of social problems in SCTs – and especially so for males with XYY.

The potential mechanisms for a prominent age-related worsening of social problems in XYY can be considered at multiple levels. First, there are behavioral considerations that may further contribute to social problems. For example, the XYY karyotype has been reliably linked to language delays and impairment, to an even greater extent than those with XXY [40]. Lower IQ scores and difficulties in verbal comprehension, working memory, and processing speed may also contribute to difficulties in social functioning [9]. These difficulties could lead to fewer opportunities for social learning across settings because of social isolation and cumulative effects of combined co-impairments of language and attention. These differences may then be exacerbated over time due to a failure to catch up to social expectations. Age-varying social impairments in XYY syndrome may also be reflected in Y-chromosome effects on neurobiological systems implicated in social processing. Specifically, additional Y chromosome dosage is known to influence cortical anatomy in brain systems that subserve social cognition and decision-making [41, 42]. We currently lack studies of age-related SCT effects on human brain organization, but there is preliminary evidence that sex chromosome dosage effects on mammalian brain anatomy can vary with age [43]. Such age-modulation of sex chromosome dosage effects on brain organization could theoretically reflect age-dependence of known SCT effects on genome function [44, 45].

Second, although age-related variation is most prominent for social problems in XYY, we also see hints of age effects on other domains in other SCTs, most notably age-related increases of internalizing problems in XXX. The lack of statistical significance of this association, despite its magnitude being similar to that of age effects on social problems in XYY syndrome, should be interpreted in light of the differences in sample size between XXX and XYY cohorts. Future work should increase sample size and balance representation of different SCTs to test for karyotype specificity of age influences on psychopathology. It is notable that just as social problems are both more severe in XYY and more prone to age-related exacerbation, internalizing problems are also relatively pronounced in XXX (**Supplementary Table 2**). Thus, a speculative, but intriguing possibility is that those domains of psychopathology showing pronounced impacts in a given SCT are also the ones that are most prone to worsening with increasing age.

Third, we noted a qualitative heterogeneity of age-related variation of psychopathology across measured domains in SCT groups as compared to euploidic controls. It is notable for example that there is a marked, though non-significant, decrease in rule breaking, aggressive, and overall externalizing behaviors in XYY. Additionally, while we see an increase in internalizing problems in XXX with age, internalizing problems show a declining trajectory in XXY. Hence, more research is needed in larger samples to parse the complex relationship between symptoms, karyotypes, and symptom change over time.

Fourth, the dataset we used to estimate age-related variation of psychopathology in SCT enabled us to re-visit a question in our previous work investigating mean differences in psychopathology scores in youth with sex chromosome aneuploidies. Our findings reinforce the results from this earlier work, in an expanded sample size [10]. Present analyses confirmed that XYY males tend to experience more social, thought, and attention problems than XXY [10]. Novel findings from analyses in the expanded sample of our present study versus our prior study [10] are that: (i) Attention Problems were statistically significantly higher in both XXX and XYY as compared XXY (whereas a significant difference was only observed between XYY and XXY in our prior work); (ii) XYY showed significantly greater mean T-scores than XXY for Somatic Complaints, Rule-Breaking Behavior, and Internalizing, and; (iii) XYY showed significantly greater mean T-scores than XXY and XXX for Social Problems, Thought Problems, Aggressive Behavior, Externalizing, and Total Problems (previously a significant difference was only only observed between XYY and XXY for the Social and Thought scales). We attribute the additional significant findings detailed above primarily to increased sample sizes considering that mean CBCL T-scores were similar between Rau et al. [10] and the current study. The largest difference in CBCL mean score between studies was 4.4 T-score units, less than half of a standard deviation. Importantly these statements about the main effects of SCT group on severity of psychopathology exist within the context of age-related variation, which is the primary focus of the study.

Our findings must be considered in light of several potential caveats and limitations. First, a primary issue with the current data is the unbalanced sample size across karyotypes. While a goal in the field has been to increase the sample size of studies featuring the rare sex chromosome aneuploidy disorders, maintaining equality across karyotypes is similarly important, as imbalance impacts statistical power and therefore the conclusions that can be drawn about one karyotype over another. Additionally, we used only one measure of psychopathology, but our understanding of an individual’s experience would be richer if we used finer-grained measures of psychopathology, both in the depth and breadth of symptoms measured, and varied the type of report collected (i.e., self-versus parent report). The present study features a cross-sectional data set which prevents us from measuring within-person change, and we are also prone to potential age-related ascertainment biases influencing cross-sectionally estimated age effects. A priority for future research will be to gather longitudinal behavioral datasets in SCT that are sufficiently large to permit tests for potential non-linear age effects and modulation of age effects by variation in diverse clinical (e.g., IQ) or demographic (e.g., SES, trauma) variables.

Notwithstanding these limitations and caveats, our study provides a valuable survey of age-related variation in all three SCT syndromes across childhood and adolescence. We find that the severity of psychopathology within SCT can vary heterogeneously across different karyotypes and domains of psychopathology – a finding which carries important implications for personalization of clinical care in SCT and motivates expanded research on age-varying impacts of SCT on structure and function of the human brain.

## Data Availability

The datasets used and analyzed during the current study are available from the corresponding author on reasonable request.

## Acknowledgements

We thank the patients and their families for participating in this study, as well as the Association for X and Y Chromosome Variations (https://genetic.org) for their assistance with recruitment.

