## SUPPLEMENTARY TABLES AND FIGURES for "Age-related differences in psychopathology within sex chromosome trisomies"

| | Total | XX | XY | XXX | XXY | XYY | Group Comparisons | | |
|---|---|---|---|---|---|---|---|---|---|
| | | | | | | | Comparing all groups | SCTs vs. controls | Comparing SCT groups only |
| **Federal Race Category: N(%)** | | | | | | | | | |
| Asian | 11 (2.2) | 3 (2.0) | 3 (2.0) | 3 (6.0) | 1 (1.1) | 1 (1.7) | $\chi2$= 35.87* | $\chi2$= 34.99** | $\chi2$= 9.94 |
| Black | 34 (6.7) | 17 (11.3) | 13 (8.5) | 0 (0) | 4 (4.3) | 0 (0) | | | |
| American Indian / Alaska Native | 1 (0.2) | 0 (0) | 0 (0) | 0 (0) | 1 (1.1) | 0 (0) | | | |
| White | 424 (84) | 123 (81.5) | 120 (78.4) | 44 (88.0) | 81 (88.0) | 56 (95.0) | | | |
| More than one race | 33 (6.5) | 6 (4.0) | 17 (11.1) | 3 (6.0) | 5 (5.4) | 2 (3.4) | | | |
| Unknown / not reported | 2 (0.4) | 2 (1.3) | 0 (0) | 0 (0) | 0 (0) | 0 (0) | | | |
| **Federal Ethnicity Category: N(%)** | | | | | | | | | |
| Hispanic | 34 (6.7) | 10 (6.6) | 6 (4) | 4 (8) | 9 (9.8) | 5 (8.5) | $\chi2$= 9.23 | $\chi2$= 3.60 | $\chi2$= 2.58 |
| Non-Hispanic | 468 (92.7) | 141 (93.4) | 146 (95.4) | 46 (92) | 81 (88) | 54 (91.5) | | | |
| Unknown | 3 (0.6) | 0 (0) | 1 (0.7) | 0 (0) | 2 (2.2) | | | | |

**Supplementary Table 1 Additional demographic variables** * $p < .05$, **$p < .001$

| CBCL Scale | CBCL T-score by group, M(SD) | | | | |
|---|---|---|---|---|---|
| | XX | XY | XXX | XXY | XYY |
| anxdep | 51 (0.5) | 51.3 (0.5) | 60.3 (1.1)* | 57.9 (0.8)* | 59.7 (1.0)* |
| withdep | 51.5 (0.5) | 52.0 (0.5) | 61.7 (1.1)* | 58.9 (0.8)* | 61.3 (1.0)* |
| somatic | 52.2 (0.5) | 52.3 (0.5) | 62.6 (1.1)* | 59.2 (0.9)* | 63.3 (1.0)* |
| social | 50.6 (0.5) | 51.2 (0.5) | 63.5 (1.0)* | 60.7 (0.8)* | 68.1 (0.9)* |
| thought | 51.1 (0.5) | 51.7 (0.5) | 60.5 (1.0)* | 60.0 (0.8)* | 66.0 (1.0)* |
| attention | 50.9(0.5) | 51.4 (0.5) | 67.1 (1.1)* | 60.9 (0.8)* | 68.6 (1.0)* |
| rulebreak | 51.3 (0.4) | 51.4 (0.4) | 56.6 (0.9)* | 56.1 (0.7)* | 59.0 (0.8)* |
| aggressive | 50.7 (0.5) | 51.0 (0.5) | 58.1 (1.0)* | 56.5 (0.8)* | 62.6 (0.9)* |
| internal | 42.5 (0.8) | 43.6 (0.8) | 61.7 (1.6)* | 57.6 (1.3)* | 62.2 (1.5)* |
| external | 42.0 (0.8) | 42.6 (0.8) | 53.9 (1.6)* | 53.8 (1.2)* | 59.4 (1.5)* |
| totalprob | 40.0 (0.8) | 41.3 (0.7) | 61.4 (1.6)* | 58.7 (1.2)* | 65.8 (1.4)* |

**Supplementary Table 2 Mean CBCL T-score for each CBCL dimension in each karyotype group (estimated at mean age of the combined cohort).** Asterisk denotes Tukey post hoc adjusted *p*-value for comparison of each SCT group's CBCL T-score distribution relative to their respective gonadal control group ($p < .05$).

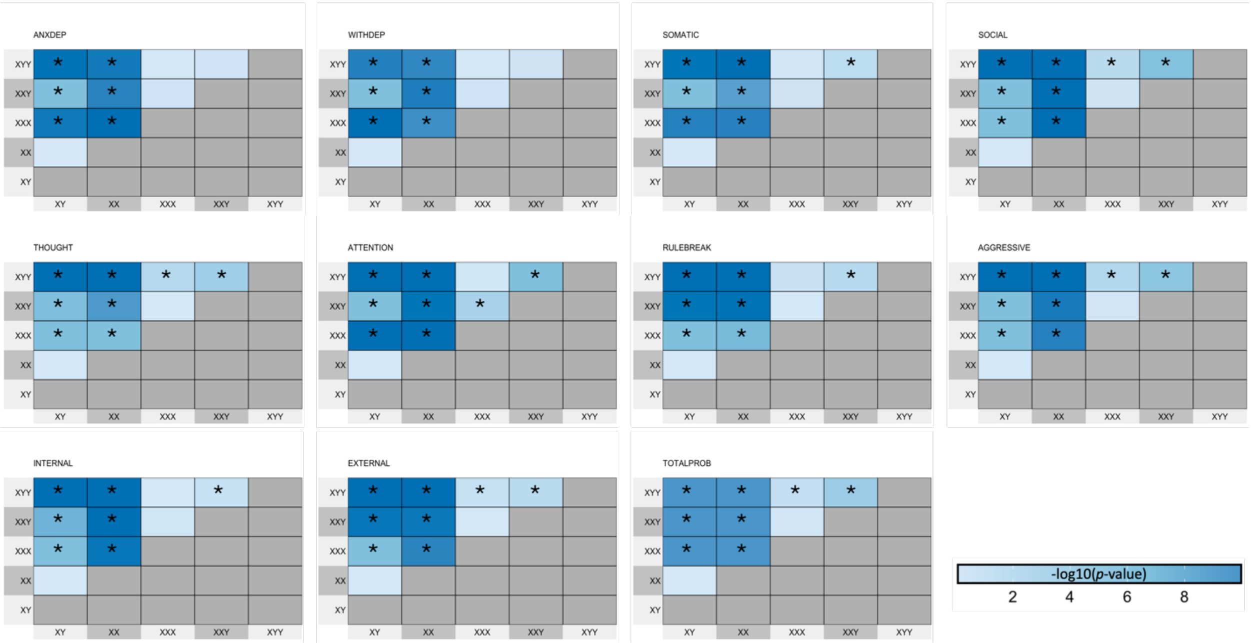


**Supplementary Fig. 1 Pairwise tests for inter-group differences in mean CBCL T-score for each CBCL dimension.** Colors encode the -log10(*p*-value) from Tukey test for each group difference. Asterisks denote statistically significant contrasts (adjusted *p*-value < .05). Directions of group differences are detailed in Supplementary Table 2.
